# Preparing for the Postpartum Medicaid Extension Evaluation in Illinois

**DOI:** 10.1101/2024.05.03.24306844

**Authors:** Arden Handler, Caitlin Meyer Krause, Kristin Rankin

## Abstract

**Background:** Given increased attention to the maternal health crisis, its heavy toll on Black birthing persons, and recognition that most pregnancy-related deaths occur in the extended postpartum period, Illinois implemented a Postpartum Medicaid Extension (PME).

**Objectives:** The purpose of this study was to use baseline data from the pre-PME period to ascertain which groups of eligible persons can be expected to experience improved outcomes as a result of PME implementation in Illinois.

**Methods:** We focused on the Well-Woman Visit (WWV) as it can be measured in the later postpartum period and is relevant for non-pregnant women. We provide baseline prevalence estimates for WWV receipt using PRAMS and BRFSS data within income strata and within combined income-insurance strata with particular attention to the income group most likely to be affected by the PME. Using multivariable binomial regression, we generate adjusted prevalence differences across income and insurance strata overall and by race/ethnicity.

**Results:** The Illinois PME has the potential to improve the receipt of well-woman care in the 138-213% FPL income group, the group most likely to be affected by the PME. The analysis also suggests that Black women in Illinois may not be the group most likely to benefit from the enhanced access to care made available by the PME without additional focused attention to their particular needs and experiences. Conclusions and Implications for Practice and Policy:

The PME is necessary but not sufficient for addressing racial/ethnic inequities in maternal health. Leveraging the opportunity that the PME provides to design and support delivery models that maximize the effects of such coverage will be essential to address the maternal health crisis in Illinois. Without extra attention to the needs and experiences of Black women, focused on the delivery of care as well as the structural determinants of health including institutional and interpersonal racism, the benefits of the PME for Black women may not be fully realized.

**Recommendations for Practice:**

1. An extensive outreach and communications campaign focused on the Postpartum Medicaid Extension in Illinois is necessary so that all birthing persons covered by Medicaid and their providers know that postpartum coverage now continues through 12 months.
2. New approaches to care are needed to maximize the benefits of the PME, including elevation of the medical care home for women’s primary/interconception care, efforts to reduce provider bias in the delivery of care, and implementation of postpartum care models such as the Two-Generation approach.
3. Development of a Performance Measurement System within Medicaid focused on the extended postpartum period is essential to ensure that Medicaid providers are aware of the PME and pay particular attention to the extended postpartum period.
4. Promotion of enhanced postpartum care packages that include reimbursement for services to address the structural/social determinants of health is essential.

## Introduction

Medicaid’s role in providing coverage for women’s reproductive and pregnancy health has expanded greatly since its enactment in 1965.^1^ Changes beginning in the mid-1980s ultimately led to the decoupling of cash assistance receipt from Medicaid coverage during pregnancy through 60 days postpartum. Expansions continued with the mid-1990s Family Planning Medicaid expansion,^2^ and the provision of Medicaid coverage to low-income non-pregnant adults through the Affordable Care Act (2014).

Given increased attention to the maternal health crisis,^3,4^ its heavy toll on Black birthing persons, and recognizing most pregnancy-related deaths occur in the extended postpartum period,^3,5^ the latest Medicaid expansion extends Medicaid coverage beyond 60 days to 12 months postpartum. While states could implement a postpartum Medicaid extension with their own funds or a 1115 waiver,^6^ the American Rescue Plan Act passed during the COVID-19 pandemic allowed states to temporarily extend Medicaid coverage through 12 months postpartum using a State Plan Amendment (SPA), with significant advantages over waivers.^6–8^ Postpartum Medicaid Extension (PME) via a SPA became a permanent option through the Omnibus Reconciliation Act of 2022;^9^ evaluations of PMEs are necessary to determine their effect on improving maternal/women’s health.

In April 2021, Illinois became the first state to have a 1115 waiver approved by the Centers for Medicare and Medicaid Services (CMS), providing full benefit Medicaid coverage through 12 months postpartum (effective through 12-25). In September 2021, CMS approved a Title XXI (CHIP) Health Services Initiative (HSI) SPA that enabled Illinois to extend coverage to immigrants who did not qualify under the 1115 waiver. These postpartum extension authorities were estimated to cover almost 10,000 persons annually. Although Illinois maintained the distinction of having the first approved PME, this waiver never went into effect due to the COVID-19 Public Health Emergency (PHE) in which coverage automatically continued for all Medicaid recipients. During this period, a SPA was obtained to replace the 1115 waiver. When the PHE ended in May 2023, Illinois’ PME SPA (rather than waiver) went into effect.

Evaluations of Medicaid expansions such as the PME face multiple challenges. Separate from expansion implementation concerns, a major methodological issue is that Medicaid recipients in the post-expansion period are different from recipients in the pre-expansion period. Newly eligible persons are only identifiable in Medicaid claims data, post-expansion. Therefore, evaluations must account for demographic changes from pre-to post-expansion in the composition of the Medicaid covered group and for secular trends in outcomes or time-varying factors that would have occurred with or without the expansion.

Much can be learned from previous efforts to evaluate Medicaid expansions. Scholars have used a variety of approaches to account for differences in the Medicaid recipient groups pre- and post-expansion. The general strategy is to compare trends over time at the population level in selected outcomes before and after the expansion (with or without a comparison group) within the same^10^ or different geographic jurisdictions.^11,12^ In this strategy there is an expectation of discontinuity in trends from the pre-to post-expansion periods.^10–12^ Difference-in-differences methods are commonly employed.^10,12–16^

Drilling down to **specific approaches**, researchers often identify *a priori* a demographic group most likely to be affected by the Medicaid expansion and examine outcomes within that group pre- and post-expansion.^17^ Another strategy is to examine changes in outcomes in a non-privately insured group (Medicaid and uninsured combined) pre- and post-expansion, acknowledging that the proportion of Medicaid-covered individuals will increase and the proportion of uninsured individuals will decrease between periods.^18^ The latter addresses potential selection bias that occurs if participants among the expansion-eligible group are “healthier” than non-participants.

A translation of these two strategies into approaches for evaluating the Illinois PME is described below:

### Strategy 1

Using a non-Medicaid claims dataset that has <u>income information</u> (e.g., Behavioral Risk Factor Surveillance System, BRFSS), examine change over time (pre-versus post-PME) in outcomes within income strata with the *a priori* expectation for the greatest positive change in the group most affected by the PME (i.e., the 138%-213% FPL group).

### Strategy 2

Using a non-Medicaid claims dataset that has both <u>income and insurance type information (</u>e.g., Pregnancy Risk Assessment Monitoring System, PRAMS), compare outcomes in the pre-versus post-period for the 138%-213% FPL group without private insurance. Focusing on the same demographic group in both periods controls for selection bias; *a priori*, health care utilization and health status are expected to improve in the “without private insurance group,” relative to the privately insured group, post-extension.

The purpose of this study was to use baseline data from the pre-PME period to ascertain which groups of eligible persons can be expected to experience improved outcomes as a result of PME implementation in Illinois. We focused on the outcome of receiving a **Well-Woman Visit (WWV)**, as it can be measured in the later postpartum period, is relevant for non-pregnant women (those beyond 60 days postpartum), and is available in both the PRAMS and BRFSS datasets. Thus, we produced baseline prevalence estimates for WWV receipt within income strata (Strategy 1 pre-PME estimates) and within combined income-insurance strata (Strategy 2 pre-PME estimates) and generated adjusted prevalence differences (aPD) to compare the prevalence of the WWV across income and insurance strata, overall and by race/ethnicity. Finally, we highlight the benefits and challenges of using PRAMS and BRFSS to evaluate the PME.

## Materials and Methods

PRAMS and BRFSS were used to generate baseline measures for Strategy 1; only PRAMS was used for Strategy 2 baseline measures as Illinois BRFSS does not have a variable for insurance type. Of note, BRFSS does not provide information about pregnancy status in the year before the WWV, while in PRAMS, WWV information is collected with respect to the 12 months prior to the index pregnancy. As such, when referring to BRFSS analyses we use the term “women”; when referring to PRAMS analyses, we use the terms “women” or “birthing persons”.

### BRFSS Analysis

#### Strategy 1 Baseline

Illinois 2018-2019 BRFSS^19^ data were used to examine women’s WWV experiences prior to the PHE and the PME. The sample was restricted to women of reproductive age (WRA), ages 18-44 years (n=1908), who were not missing information on income and household size (missing n=184) or WWV attendance (missing n=6), resulting in a sample size of 1718 individuals.

### Exposure

Household income was collected as a categorical variable and category midpoints were assigned for each woman’s household income.^20^ Using household income and size, income as a percentage of the federal poverty level (FPL) was calculated and categorized as follows: <138% FPL, 138-213% FPL, and >213% FPL per Medicaid eligibility for non-pregnant (<138% FPL) and pregnant (≤213% FPL) persons in IL. The PME enables women in the 138-213% FPL group to remain on Illinois Medicaid for one year postpartum.

### Outcome

Our outcome of interest was receipt of a WWV in the past 12 months. This information comes from the question “About how long has it been since you last visited a doctor for a routine checkup?” which was recoded as binary. Those who selected “within the past year (anytime less than 12 months ago)” were coded as “yes” with responses of any longer time or “never” coded as “no”.

### Covariates

Covariates of interest included race and ethnicity (Non-Hispanic [NH] White, NH Black, Hispanic, NH other), age group (18-24, 25-34, 35+), marital status (married, unmarried), education (less than high school, high school diploma/GED, some college, bachelor’s degree or higher), and survey language (English, Spanish).

### Statistical Analyses

BRFSS data were weighted to produce estimates representative of WRA in Illinois. The distribution of demographic characteristics was compared across income groups using chi-square tests. We estimated pre-extension WWV prevalence and differences by income group and covariates. Using multivariable binomial regression, aPDs and 95% confidence intervals (CI) for the associations between income group and receipt of a WWV were generated overall and for each racial/ethnic group.

### PRAMS Analysis

#### Strategy 1 Baseline

Illinois PRAMS^21^ Phase 8 data included 5147 respondents from 2016-2019. The eligible sample was restricted to individuals with ≥1 prior livebirth (n=3193) since the postpartum period associated with a prior birth (i.e., preconception period for the index birth in PRAMS) is of interest for this study. After restricting to those with complete information to calculate their income as a percent of the FPL, the total sample size was 2905. PRAMS is a complex sample survey; data were weighted to produce estimates representative of multiparous persons with a recent live birth in Illinois.

### Exposure

Household income in the 12 months before the index infant was born was collected as a categorical variable and category midpoints were assigned for each birthing person’s household income. Income as a percentage of the FPL was calculated by combining household income and household size prior to the birth of the index infant.^22^ Cutoffs for the three-category income variable were the same as for the BRFSS analysis.

### Outcome

To “mimic” well-woman care utilization in the extended postpartum period to the extent possible, we restricted the analysis to multiparous PRAMS participants and used the following survey question: “What type of health care visit did you have in the 12 months before you got pregnant with your new baby?” A combined dichotomous *prepregnancy check-up* variable was coded “yes” for affirmative responses to any one of the following non-mutually exclusive options: responses: “Regular checkup at my family doctor’s office”, “Regular checkup at my OB/GYN’s office”, and “Visit for family planning or birth control.”

### Covariates

Categorical insurance type (private insurance, Medicaid, other Insurance, uninsured) was measured for the month prior to pregnancy. Additional demographic covariates included: maternal age (≤24, 25-29, 30-34, ≥35), parity (1-2 prior live births, 3 or more prior live births), maternal race/ethnicity, education, and marital status. The categories for the last three variables are consistent with the BRFSS analysis.

### Statistical Analysis

The distribution of demographic characteristics and the prevalence of the combined prepregnancy check-up outcome were generated for each income group. Adjusted PDs and 95% CIs were generated from multivariable binomial regression models using >213% FPL as the common referent group to assess the relationship between income group and receipt of a combined prepregnancy check-up, overall and within strata of race/ethnicity.

### Strategy 2 Baseline

To understand the effect that the PME may have on newly eligible postpartum individuals, this analysis was restricted to the 138-213% FPL group (n=418) and adds **insurance type** information available in the PRAMS dataset to the income information utilized in Strategy 1 Baseline. Individuals missing data on prepregnancy insurance type (n=11) or with “other” insurance (n=10) were excluded for a final sample size of 397 postpartum individuals.

### Exposure

Self-reported insurance type during the month prior to the index pregnancy was the exposure variable. A binary variable was created from the categories reported: Medicaid/uninsured or private insurance.

### Outcome and Covariates

The outcome and covariates are the same as in Strategy 1 Baseline.

### Statistical Analysis

To carry out the baseline analysis for Strategy 2 within the 138-213% FPL income stratum, the Medicaid/Uninsured group was compared to those who had Private insurance prior to the index pregnancy. Demographic characteristics were generated and the prevalence of the combined prepregnancy check-up outcome was estimated by insurance type. Adjusted PDs comparing the prevalence of receiving a prepregnancy check-up for the Medicaid/Uninsured group to the Private insurance group were generated overall and within racial/ethnic strata using multivariable binomial regression models.

All analyses for both strategies used the complex sample survey procedures in SAS 9.4 and Stata 13.1 to account for the sampling design and weighting of BRFSS and PRAMS.

## Results

### Strategy 1 Baseline

#### BRFSS

Based on BRFSS data from 2018-2019, the population estimates for the percent of WRA in the <138%, 138-213%, and >213% FPL income groups were 36.6%, 16.9%, and 46.5%, respectively. Most WRA with incomes between 138-213% FPL were 25-34 years of age, NH White, unmarried, and had some college education. Compared to the >213% FPL group, distributions of each characteristic were significantly different for the <138% (p<0.001) and 138-213% FPL (p<0.05) groups (Table 1).

**Table 1:** Distribution of Demographic Characteristics by Income Group (Strategy 1 Baseline) among Women of Reproductive Age (18-44), Illinois BRFSS 2018-2019.

|  | <138% FPL<br>(unweighted n=611) | 138-213% FPL<br>(unweighted n=265) | >213% FPL<br>(unweighted n=842) |
| --- | --- | --- | --- |
|  | Weighted%<br>(95% CI) | Weighted%<br>(95% CI) | Weighted%<br>(95% CI) |
| **Age Group** |  |  |  |
| 18-24 | 27.8<br>(23.3, 32.3) | 27.8<br>(20.9, 34.8) | 18.0<br>(14.4, 21.5) |
| 25-34 | 38.4<br>(33.8, 43.1) | 37.6<br>(30.8, 44.4) | 38.6<br>(34.7, 42.4) |
| 35-44 | 33.7<br>(29.5, 37.9) | 34.6<br>(28.2, 41.1) | 43.5<br>(39.6, 47.3) |
| **Race/Ethnicity** |  |  |  |
| White, NH | 35.3<br>(30.7, 40.0) | 54.0<br>(46.9, 61.1) | 70.8<br>(67.2, 74.4) |
| Black, NH | 18.2<br>(14.5, 21.8) | 24.4<br>(18.1, 30.8) | 12.9<br>(9.9, 15.9) |
| Hispanic | 36.7<br>(32.5, 41.0) | 15.6<br>(10.6, 20.5) | 7.0<br>(5.5, 8.6) |
| Other/Multiracial,<br>NH | 9.8<br>(6.3, 13.3) | 6.0<br>(2.9, 9.1) | 9.2<br>(6.9, 11.6) |
| **Education** |  |  |  |
| <HS | 26.4<br>(22.0, 30.9) | 5.1<br>(0.9, 9.3) | 1.2<br>(0.0, 3.1) |
| HS Diploma or GED | 34.2<br>(29.7, 38.7) | 24.8<br>(18.7, 30.9) | 10.4<br>(7.8, 13.1) |
| Some college | 28.2<br>(23.9, 32.5) | 48.4<br>(41.3, 55.5) | 29.9<br>(25.9, 33.9) |
| Bachelor's or higher | 11.1<br>(8.8, 13.5) | 21.7<br>(16.9, 26.5) | 58.4<br>(54.3, 62.3) |
| **Marital Status** |  |  |  |
| Married | 24.8<br>(20.8, 28.9) | 38.4<br>(31.5, 45.2) | 55.3<br>(51.3, 59.3) |
| Unmarried | 75.2<br>(71.1, 79.2) | 61.6<br>(54.8, 68.5) | 44.7<br>(40.7, 48.7) |
| **Survey Language** |  |  |  |
| English | 73.8<br>(70.0, 77.6) | 95.9<br>(93.8, 98.0) | 99.7<br>(99.4, 100.0) |
| Spanish | 26.2<br>(22.4, 30.0) | 4.1<br>(2.0, 6.2) | 0.3<br>(0.0, 0.6) |
| **Insurance Coverage** |  |  |  |
| Yes | 70.4<br>(66.3, 74.5) | 81.1<br>(75.1, 87.1) | 96.0<br>(94.5, 97.6) |
| No | 29.6<br>(25.5, 33.7) | 18.9<br>(12.9, 24.9) | 4.0<br>(2.4, 5.5) |

Note: All variables have significantly different distributions for the <138% vs. >213% FPL groups (p<0.001) and for the 138-213% vs. >213% FPL groups (p<0.05).

The self-reported prevalence of receiving a WWV in the past 12 months for WRA with income levels of <138%, 138-213%, and >213% FPL was 73.8%, 72.1%, and 79.0%, respectively (Table 2). After adjusting for covariates, WRA with incomes 138-213% FPL had a WWV prevalence 7.5 percentage points lower (95% CI: -14.3, -0.7) than WRA with incomes >213% FPL.

**Table 2.** Prevalence of Receipt of a Well-Woman Visit by Income Group (Strategy 1 Baseline) among Women of Reproductive Age (18-44), with Adjusted Prevalence Differences, Overall and Stratified by Race/Ethnicity, Illinois BRFSS 2018-2019.

|  | **<138% FPL**<br>**Weighted%**<br>**(95% CI)** | **138-213% FPL**<br>**Weighted%**<br>**(95% CI)** | **>213% FPL**<br>**Weighted%**<br>**(95% CI)** | **aPD<sup>a</sup> <138%**<br>**vs >213%**<br>**(95% CI)** | **aPD<sup>a</sup> 138-213%**<br>**vs >213%**<br>**(95% CI)** |
| --- | --- | --- | --- | --- | --- |
| Overall<sup>b</sup> (n=1718) | 73.8<br>(69.7, 77.9) | 72.1<br>(65.7, 78.5) | 79.0<br>(76.0, 82.1) | -3.7%<br>(-9.2%, 1.7%) | -7.5%<br>(-14.3%, -0.7%) |
| NH White (n=895) | 69.5<br>(61.5, 77.4) | 64.6<br>(55.6, 73.6) | 77.0<br>(73.2, 80.8) | -8.0%<br>(-17.0%, 1.0%) | -12.9%<br>(-22.8%, -3.0%) |
| NH Black<br>(n=243) | 91.2<br>(85.3, 97.0) | 84.2<br>(73.9, 94.5) | 86.8<br>(79.9, 93.7) | 5.4%<br>(-5.1%, 15.9%) | -1.2%<br>(-13.1%, 10.8%) |
| Hispanic/ Latina<br>(n=440) | 66.7<br>(60.4, 73.0) | 70.2<br>(50.9, 89.4) | 79.8<br>(70.8, 88.7) | -9.5%<br>(-21.6%,<br>-2.6%) | -10.8%<br>(-28.3%, 6.6%) |

aPD=Adjusted Prevalence Difference; NH=Non-Hispanic/Latina
<sup>a</sup>Adjusted for age, race/ethnicity, and education for overall estimates, and age and education for the race/ethnicity stratified estimates.
<sup>b</sup>Overall results include non-Hispanic women of other races, but they are not shown as a race/ethnicity stratum due to insufficient sample size.

Within each income group, the self-reported prevalence of receiving a WWV was highest for NH Black compared to NH White and Hispanic women. Among NH White WRA, compared to women with incomes >213% FPL, the prevalence of receiving a WWV was 12.9 percentage points lower for those with incomes 138-213% FPL (95% CI: -22.8, -3.0), after adjusting for age and education. In adjusted analysis for Hispanic WRA, those with incomes <138% FPL had a WWV prevalence 9.5 percentage points lower (95% CI: -21.6, -2.6) than those with incomes >213% FPL. All other adjusted associations, including those among NH Black women, had 95% CIs that included the null.

#### PRAMS

Using 2016-2019 PRAMS data, population estimates for the percent of birthing persons with ≥1 previous livebirth in the <138%, 138-213%, and >213% FPL income groups were 39.3%, 14.2%, and 46.5%, respectively. Distributions were generated for all demographic variables by income group (Table 3, Strategy 1). Most birthing persons with incomes 138-213% FPL were 25-29 years, NH White, married, had some college education, and private insurance. Distributions of each characteristic were significantly different when comparing the <138% (p<0.0001) and 138-213% FPL (p<0.01) groups to those with income >213% FPL.

**Table 3:** Distribution of Demographic Characteristics by Income Group (Strategy 1 Baseline) and Insurance Type in the 138-213% FPL Group (Strategy 2 Baseline) among Postpartum Persons with One or More Prior Livebirths, Illinois PRAMS 2016-2019.

|  | **Strategy 1 Baseline: Income Group<sup>a</sup>** |  |  |  | **Strategy 2 Baseline: Prepregnancy Insurance type among 138-213% FPL<sup>b</sup>** |  |
| --- | --- | --- | --- | --- | --- | --- |
|  | **<138% FPL**<br>**(unweighted n=1144)**<br>**Weighted%**<br>**(95% CI)** | **138-213% FPL**<br>**(unweighted n=418)**<br>**Weighted%**<br>**(95% CI)** | **>213% FPL**<br>**(unweighted n=1343)**<br>**Weighted%**<br>**(95% CI)** |  | **Medicaid/Uninsured**<br>**(unweighted n=229)**<br>**Weighted%**<br>**(95% CI)** | **Private**<br>**(unweighted n=168)**<br>**Weighted%**<br>**(95% CI)** |
| **Age** |  |  |  |  |  |  |
| ≤24 | 24.0<br>(21.1, 26.9) | 14.7<br>(10.7, 18.8) | 3.0<br>(2.0, 4.1) |  | 20.4<br>(14.1, 26.7) | 6.8<br>(2.2, 11.4) |
| 25-29 | 32.2<br>(29.2, 35.3) | 36.1<br>(30.9, 41.3) | 20.0<br>(17.7, 22.3) |  | 41.2<br>(33.8, 48.6) | 29.7<br>(22.0, 37.3) |
| 30-34 | 25.8<br>(23.0, 28.7) | 27.7<br>(23.0, 32.3) | 43.3<br>(40.5, 46.1) |  | 21.6<br>(15.9, 27.3) | 33.9<br>(26.2, 41.7) |
| ≥35 | 17.9<br>(15.5, 20.3) | 21.5<br>(17.2, 25.8) | 33.7<br>(31.0, 36.4) |  | 16.8<br>(11.5, 22.1) | 29.6<br>(22.1, 37.2) |
| **Race/Ethnicity** |  |  |  |  |  |  |
| NH White | 34.4<br>(31.2, 37.6) | 45.0<br>(39.6, 50.4) | 77.5<br>(75.1, 79.9) |  | 45.4<br>(38.0, 52.8) | 47.3<br>(38.9, 55.7) |
| NH Black | 28.4<br>(25.5, 31.4) | 18.9<br>(14.6, 23.1) | 5.0<br>(3.8, 6.3) |  | 19.3<br>(13.6, 25.0) | 20.4<br>(13.3, 27.4) |
| Hispanic/Latina | 34.1<br>(30.9, 37.3) | 32.0<br>(26.8, 37.2) | 9.6<br>(7.9, 11.4) |  | 32.1<br>(25.0, 39.3) | 28.6<br>(20.8, 36.4) |
| NH Other | 3.0<br>(1.9, 4.1) | 4.1<br>(2.1, 6.2) | 7.8<br>(6.3, 9.4) |  | 3.2<br>(0.9, 5.5) | 3.7<br>(0.6, 6.9) |
| **Parity** |  |  |  |  |  |  |
| 1-2 prior live births | 70.4<br>(67.5, 73.4) | 80.4<br>(76.2, 84.6) | 92.2<br>(90.7, 93.7) |  | 80.6<br>(74.9, 86.2) | 82.4<br>(76.1, 88.8) |
| 3 or more prior live births | 29.6<br>(32.5, 64.4) | 19.6<br>(14.5, 23.8) | 7.8<br>(6.3, 9.3) |  | 19.4<br>(74.9, 86.2) | 17.6<br>(11.2, 23.9) |
| **Education** |  |  |  |  |  |  |
| <HS | 21.8<br>(19.0, 24.6) | 11.2<br>(7.8, 14.6) | 1.7<br>(0.9, 2.4) |  | 14.1<br>(9.0, 19.2) | 5.6<br>(1.8, 9.5) |
| HS Diploma or GED | 39.9<br>(36.6, 43.2) | 33.8<br>(28.6, 39.1) | 6.8<br>(5.2, 8.3) |  | 37.8<br>(30.4, 45.1) | 27.5<br>(19.8, 35.1) |
| Some college | 31.4<br>(28.3, 34.5) | 37.8<br>(32.6, 42.9) | 20.2<br>(17.9, 22.6) |  | 37.0<br>(29.9, 44.1) | 40.1<br>(31.9, 48.2) |
| Bachelor's or higher | 6.9 | 17.2 | 71.3 |  | 11.1 | 26.8 |
|  | **<138% FPL**<br>**(unweighted n=1144)**<br>**Weighted%**<br>**(95% CI)** | **138-213% FPL**<br>**(unweighted n=418)**<br>**Weighted%**<br>**(95% CI)** | **>213% FPL**<br>**(unweighted n=1343)**<br>**Weighted%**<br>**(95% CI)** |  | **Medicaid/Uninsured**<br>**(unweighted n=229)**<br>**Weighted%**<br>**(95% CI)** | **Private**<br>**(unweighted n=168)**<br>**Weighted%**<br>**(95% CI)** |
|  | (5.3, 8.5) | (13.3, 21.1) | (68.7, 74.0) |  | (6.6, 15.5) | (19.6, 34.1) |
| **Marital Status** |  |  |  |  |  |  |
| Married | 34.4<br>(31.3, 37.5) | 59.9<br>(54.6, 65.3) | 90.9<br>(89.1, 92.6) |  | 53.0<br>(45.6, 60.5) | 66.1<br>(58.0, 74.2) |
| Unmarried | 65.6<br>(62.5, 68.7) | 40.1<br>(34.7, 45.4) | 9.1<br>(7.4, 10.9) |  | 47.0<br>(39.5, 54.4) | 33.9<br>(25.8, 42.0) |
| **Insurance type – prior to pregnancy** |  |  |  |  |  |  |
| Private Insurance | 12.6<br>(10.4, 14.8) | 41.9<br>(36.5, 47.2) | 92.5<br>(91.0, 94.0) |  | 0.0 | 100.0<br>(100.0, 100.0) |
| Medicaid | 65.1<br>(61.9, 68.2) | 37.3<br>(32.0, 42.6) | 3.8<br>(2.6, 4.9) |  | 67.3<br>(60.3, 74.3) | 0.0 |
| Other | 2.0<br>(1.0, 2.9) | 2.7<br>(1.0, 4.4) | 1.7<br>(1.0, 2.4) |  | 0.0 | 0.0 |
| Uninsured | 20.3<br>(17.6, 23.0) | 18.2<br>(13.9, 22.4) | 2.0<br>(1.2, 2.8) |  | 32.7<br>(25.7, 39.7) | 0.0 |

<sup>a</sup>All variables have significantly different distributions for the <138% (p<0.0001) and the 138-213% FPL groups (p<0.01) vs. the >213% FPL group.
<sup>b</sup>All variables have significantly different distributions for the Medicaid/Uninsured vs. Private groups (p<0.02), except for race/ethnicity, and parity.

The self-reported prevalence of receiving a prepregnancy checkup for multiparous birthing persons with income levels of <138%, 138-213%, and >213% FPL was 48.4%, 53.8% and 72.8%, respectively (Table 4, Strategy 1). Compared to birthing persons with incomes >213% FPL, those with incomes <138% FPL had a prepregnancy checkup prevalence 11percentage points lower (95% CI: -16.7, -5.4), and those in the 138-213% FPL group had a prevalence 9 percentage points lower (95% CI: -15.7, -2.9), after adjusting for covariates.

**Table 4.** Prevalence of Receipt of a Combined Prepregnancy Checkup by Income Group (Strategy 1 Baseline) and by Insurance Type in the 138-213% FPL Group (Strategy 2 Baseline) among Postpartum Persons with One or More Prior Livebirths, with Adjusted Prevalence Differences, Overall and Stratified by Race/Ethnicity, Illinois PRAMS 2016-2019.

|  | <138% FPL<br>Weighted%<br>(95% CI) | 138-213%<br>FPL<br>Weighted%<br>(95% CI) | >213% FPL<br>Weighted%<br>(95% CI) | aPD<sup>a</sup><br><138% vs >213%<br>(95% CI) | aPD<sup>a</sup><br>138-213% vs >213%<br>(95% CI) |
| --- | --- | --- | --- | --- | --- |
| **Strategy 1 Baseline: By Income Group** |  |  |  |  |  |
| Overall<sup>b</sup><br>(n=2,902) | 48.4<br>(45.1, 51.7) | 53.8<br>(48.5, 59.2) | 72.8<br>(70.3, 75.4) | -11.0%<br>(-16.7%, -5.4%) | -9.3%<br>(-15.7%, -2.9%) |
| NH White<br>(n=1,558) | 47.6<br>(41.9, 53.3) | 56.4<br>(48.5, 54.3) | 76.2<br>(73.4, 79.0) | -16.0%<br>(-23.8%, -8.1%) | -11.6%<br>(-20.3%, -2.9%) |
| NH Black<br>(n=517) | 60.4<br>(54.3, 66.4) | 63.1<br>(51.0, 75.3) | 66.0<br>(53.7, 78.4) | 6.5%<br>(-8.7%, 21.7%) | 5.5%<br>(-11.9%, 22.8%) |
| Hispanic/<br>Latina (n=586) | 39.1<br>(33.4, 44.8) | 46.8<br>(36.8, 56.7) | 63.3<br>(54.2, 72.5) | -6.9%<br>(-21.3%, 7.4%) | -2.9%<br>(-18.7%, 12.9%) |
| **Strategy 2 Baseline: By Insurance Type in 138-213% FPL Group Only** |  |  |  |  |  |
|  | Medicaid/Uninsured<br>Weighted%<br>(95% CI) | Private<br>Weighted%<br>(95% CI) | aPD<sup>c</sup> Medicaid/Uninsured vs. Private<br>(95% CI) |  |  |
| Overall<sup>b</sup><br>(n=397) | 52.9<br>(45.5, 60.3) | 58.2<br>(50.0, 66.4) | -1.7%<br>(-13.2%, 9.9%) |  |  |
| NH White<br>(n=175) | 50.7<br>(39.8, 61.6) | 65.5<br>(54.3, 76.7) | -12.3%<br>(-28.7%, 4.1%) |  |  |
| NH Black<br>(n=90) | 75.7<br>(61.6, 89.9) | 46.0<br>(26.1, 65.1) | 28.8%<br>(4.2%, 53.3%) |  |  |
| Hispanic/Latina<br>(n=108) | 46.6<br>(32.8, 60.4) | 54.8<br>(38.6, 71.0) | -1.8%<br>(-24.0%, 20.5%) |  |  |

aPD=Adjusted Prevalence Difference; NH=Non-Hispanic/Latina
<sup>a</sup>Adjusted for maternal age, race/ethnicity, and education for overall estimates and age and education for the race/ethnicity stratified models.
<sup>b</sup>Overall results include NH women of other races, but they are not shown as a race/ethnicity stratum due to insufficient sample size.
<sup>c</sup>Adjusted for maternal age, race/ethnicity, and education for overall estimates and age and education for the race/ethnicity stratified models.

In the two lowest income groups, NH Black multiparous birthing persons had the highest self-reported prevalence of receiving a prepregnancy check-up. Among NH White multiparous birthing persons, those in the <138% income group had a 16-percentage point lower adjusted prevalence (95% CI: -23.8, -8.1), and those in the 138-213% FPL group had a 12-percentage point lower adjusted prevalence (95% CI: -20.3, -2.9) of receiving a prepregnancy checkup compared to the >213% FPL income group. The 95% CIs for the adjusted PDs between income groups for NH Black and Hispanic multiparous birthing persons all included the null (Table 4, Strategy 1).

### Strategy 2 Baseline

#### PRAMS

Overall, among multiparous birthing persons with incomes 138-213% FPL, 38.3% reported having Medicaid coverage prior to pregnancy and 18.7% were uninsured; forty-three percent reported being privately insured. The majority of those in the Medicaid/uninsured group were 25-29 years old, had at least a high school diploma or GED, and were married. Those with private insurance were mainly 30-34 years, had some college education, and were married (Table 3, Strategy 2). Distributions were significantly different by insurance type for age, education, and marital status (p<0.02).

Among the focal population, the self-reported prevalence of receiving a prepregnancy checkup in the 12 months prior to the index pregnancy for the Medicaid/uninsured and privately insured groups was 52.9% and 58.2%, respectively (Table 4, Strategy 2). The aPD comparing the two insurance groups had a 95% CI that included the null.

NH Blacks in the Medicaid/uninsured group prior to pregnancy had the highest self-reported prevalence of receiving a prepregnancy check-up among all racial/ethnic and insurance groups, 28.8 percentage points higher in adjusted analysis (95% CI: 4.2, 53.3) than the prevalence among their counterparts with private insurance. Adjusted PDs between the Medicaid/Uninsured and Private groups were not significantly different for NH Whites or Hispanics.

## Discussion

Generating baseline results for the evaluation of the PME provides insight into the groups most likely to benefit and offers an opportunity to estimate the extent of change expected. Additionally, examining baseline results can foreshadow where additional effort may be needed to ensure that the PME results in the maximum benefit for postpartum persons and leads to declines in maternal health inequities between population groups.

Based on the outcome that was the focus of this analysis, the PME in Illinois has the potential to increase the use of **well-woman care** prior to pregnancy in the 138-213% FPL group, those eligible for the PME. Based on BRFSS data, women in this income group have the lowest prevalence of receiving a WWV in the past year, an estimate similar to that of the <138% FPL group. Based on PRAMS data (Strategy 1 Baseline), multiparous birthing persons in the 138-213% FPL group have the second lowest prevalence of a prepregnancy checkup in the 12 months prior to pregnancy with the lowest prevalence among those <138% FPL, suggesting substantial room for improvement for both groups. Furthermore, aPDs between the 138%-213% and >213% FPL groups overall were significant for both data sources in the Strategy 1 baseline analysis, suggesting potential to narrow the gap between women/birthing persons eligible for the PME and their higher income counterparts.

Because the PME has gained momentum as a strategy for improving maternal health,^23,24^ particularly for the Black population which is disproportionately affected by the maternal health crisis, examining the extent to which Black birthing persons are expected to benefit from the PME in Illinois is important. Whether examining the racial/ethnic-specific prevalence of the WWV (BRFSS data, Strategy 1 Baseline) or the racial/ethnic-specific prevalence of a prepregnancy check-up (PRAMS data, Strategy 1 Baseline), Black women/birthing persons have the highest rates of well-woman care in the <138% and 138-213% FPL groups in both datasets, and in the >213% FPL based on the BRFSS data. Black birthing persons with Medicaid or no insurance also have a higher pre-pregnancy checkup prevalence than those with private insurance among those in the 138-213% FPL group (PRAMS data, Strategy 2 Baseline). These results suggest there is more room for improvement in the utilization of well-woman care for Hispanic and White compared to Black women/birthing persons. As such, unless attention is paid to the content and quality of those well-woman visits, Black women/birthing persons will not fully reap the benefits of their higher rates of attendance, particularly because they face other barriers related to the structural and social determinants of health including racism and lack of respectful care.^25^

Therefore, albeit only for one potential indicator, these data suggest that although the PME has been promoted as a strategy for reducing disparities in maternal morbidity/mortality between Black and White birthing persons, it is not clear that Black birthing persons in Illinois are most likely to benefit from the increased access to care provided by the PME. This may also be because historically, Black persons are disproportionately more likely than persons of other racial/ethnic groups to already be covered by Medicaid independent of pregnancy due to a greater likelihood of low-income and disconnection from employer-based insurance.^1,26^ In fact, according to the PRAMS data used here, Black birthing persons were the group most likely to be covered by Medicaid prior to pregnancy (data not shown).

The current situation in Illinois may be similar to the experience of the Medicaid pregnancy expansions of the early 1980’s and 1990’s,^27^ which did not lead to the expected positive changes in pregnancy outcomes.^17,28^ Although these earlier expansions increased the income eligibility cutoff for Medicaid pregnancy coverage, other access barriers were not addressed.^27^ Additionally, we now recognize the earlier pregnancy Medicaid expansions did little to improve the quality of maternal care for women already enrolled in Medicaid independent of pregnancy, who were typically the highest risk of adverse pregnancy outcomes due to lower income and multiple complex health and social issues. In the earlier expansion period, without additional attention to those already covered by Medicaid when not pregnant, improvement in birth outcomes could hardly have been expected since the pregnancy Medicaid expansions were aimed at higher income, theoretically lower-risk individuals.^29^ As such, in the current context, while expanding the months of Medicaid postpartum coverage is essential for all birthing persons, particular attention should be placed on the delivery and quality of care as well as the structural determinants of health including institutional and interpersonal racism, in order for the benefits of the PME to be fully realized.

Lessons learned from earlier Medicaid expansions must be considered as the PME is rolled out in Illinois and across the nation, including that expanded coverage is not tantamount to additional systems changes needed to ensure access to and quality of care. Zephyrin and

Johnson^24^ note that providing additional months of Medicaid coverage through the PME is only one component of what must happen to improve outcomes. Because PME coverage is full benefit coverage, the opportunity to provide birthing persons benefits beyond what might be provided in a more limited expansion should be leveraged. Most importantly, an extensive outreach and communications campaign is necessary so birthing persons covered by Medicaid and their providers know that postpartum coverage continues through 12 months. Beyond this, to maximize the benefits of the PME, new approaches to care are needed, including elevation of the medical care home for women’s primary/interconception care,^30^ and postpartum care models such as the Two-Generation approach which in some manifestations provides comprehensive and holistic dyadic postpartum care.^31-32^ Promoting enhanced postpartum care packages that include reimbursement for services to address the structural/social determinants of health is also essential.^8,25^ Finally, development of a Performance Measurement System within Medicaid focused on the extended postpartum period is necessary as a way to hold providers accountable for ensuring that postpartum persons receive the care that they are entitled to during these extra months of coverage.

The main strengths of this study are the identification of an outcome likely to be affected by the PMEs and the utilization of population-based datasets with information on those most likely to be affected. However, this study also has multiple limitations. First, the focus is on only one indicator for which change is expected from pre-to post-PME, the WWV or check-up prior to pregnancy. Although a very important outcome, at best, the WWV is a “crude” measure of women’s visits and experiences in the extended postpartum period. In future analyses, other health service utilization indicators more sensitive to the PME, and health status outcomes that can be expected to improve due to the PME must be considered; the latter will need to be derived from datasets with information on health status including acute and chronic morbidities. Importantly, the eventual PME evaluation in Illinois will need to account for the fact that during the PHE, postpartum persons could not only stay on Medicaid up through 12 months postpartum but could continue their Medicaid coverage beyond 12 months postpartum. This might result in an elevation of prevalence rates of outcomes such as the WWV. Therefore, future pre-post PME comparisons may want to consider the 2022-2023 post intervention period separately from subsequent years.

More specific limitations related to the PRAMS and BRFSS datasets should be mentioned. First, the BRFSS dataset does not have recent pregnancy status information, so the well-woman check-up estimates are for all WRA, regardless of a recent pregnancy, likely diluting the results. Furthermore, the Illinois BRFSS dataset does not have information on respondents’ insurance type, preventing its use for Strategy 2. In both datasets, income as percent of the FPL is an inexact categorization because income is captured in intervals. As such, we assigned the midpoint of each interval as the income value, which could lead to exposure misclassification. As is typical for self-reported survey data, we were also missing income data for about 10% of women in both datasets. Likewise, for our racial/ethnic group stratified analyses among those with incomes 138%-213% FPL using PRAMS data, we had limited sample size, particularly for Hispanic persons, affecting our power to detect significant PDs. In addition, all prevalence estimates are based on self-report and subject to recall limitations and misidentification of preventive versus acute care visits.

Finally, in the PRAMS analyses, birthing persons may have had different insurance in the 12 months after a prior pregnancy than in the month before the index pregnancy; in addition, for the prepregnancy checkup, the 12 months prior to the birthing person’s PRAMS-associated pregnancy may not overlap with the first 12 months after their prior pregnancy. In a sensitivity analysis among those with interpregnancy intervals (IPI) of <=12 months (n=402), the percentage of birthing persons in the 138-213% FPL group with a prepregnancy checkup increased from 53.8% overall (Table 4) to 69.6%, possibly capturing the early postpartum visit as well as a later WWV. However, the sensitivity analysis also revealed that the percentage of birthing persons in the >213%FPL group with a prepregnancy checkup increased from 72.8% (Table 4) to 83.0%, suggesting that even in this restricted IPI (<=12 months) group, there is still room for women with incomes 138-213% FPL to improve relative to those >213% FPL, supporting the findings of the main analysis.

## Conclusion

Utilizing PRAMS and BRFSS data and focused on the WWV, this baseline analysis of the pre-PME period in Illinois suggests that the PME has the potential to improve the receipt of well-woman care in the extended postpartum period in the 138-213% FPL income group, those eligible for the PME. In addition, the analysis suggests that, although the PME has been promoted as a strategy for reducing inequities in maternal morbidity and mortality, it is not clear that Black birthing persons in Illinois are most likely to fully benefit without additional focused attention to the content and quality of care to meet their specific needs and experiences, especially those related to structural issues such as institutional and interpersonal racism. As such, it is important to acknowledge that the PME is necessary but not sufficient for addressing care issues in the extended postpartum period. Rather, leveraging the opportunities that extended coverage provides to design and support delivery models that maximize the effects of such coverage will be essential to address the maternal health crisis in Illinois and the US. Without the latter, the ability of the PME to improve maternal health outcomes and reduce maternal health inequities will not be fully realized.

## Data Availability

BRFSS Data available at: Behavioral Risk Factor Surveillance System (BRFSS). Centers for Disease Control and Prevention. Behavioral Risk Factor Surveillance System. Annual Survey Data. https://www.cdc.gov/brfss/annual_data/annual_data.htm.
PRAMS data available at: PRAMS data are available as an automated research file (ARF) that can be downloaded. https://www.cdc.gov/prams/prams-data/researchers.htm

## Acknowledgements

We would like to acknowledge the support of the Illinois Department of Public Health (IDPH) Pregnancy Risk Assessment Monitoring System (PRAMS) and Behavioral Risk Factor Surveillance System (BRFSS) programs for providing access to the data used in this analysis and the Maternal and Child Health Block Grant Program (Illinois Title V) for financial support of this project. The findings and conclusions in this report are those of the author(s) and do not necessarily represent the official position of the Illinois Department of Public Health.

The authors acknowledge the constructive feedback of Kay Johnson and Dr. Jennifer Kwok on earlier versions of this manuscript.

## Notes

The authors have no conflicts of interest to disclose. This research received a *Not Human Research Determination* by the UIC Office for the Protection of Research Subjects on April 28, 2023. The authors have not received any payments or services in the past 36 months from a third party that could be perceived to influence, or give the appearance of potentially influencing, the submitted work.

### Competing Interest Statement

The authors have declared no competing interest.

### Funding Statement

The Illinois Maternal and Child Health Block Grant Program of the Illinois Department of Public Health provided financial support for this project.

### Author Declarations

The University of Illinois at Chicago Office for the Protection of Research Subjects provided a Not Human Research Determination on April 28, 2023.

